# The NeoSep Severity and Recovery scores to predict mortality in hospitalized neonates and young infants with sepsis derived from the global NeoOBS observational cohort study

**DOI:** 10.1101/2022.06.21.22276677

**Authors:** Neal Russell, Wolfgang Stöhr, Aislinn Cook, James A Berkley, Bethou Adhisivam, Ramesh Agarwal, Nawshad Uddin Ahmed, Manica Balasegaram, Neema Chami, Adrie Bekker, Davide Bilardi, Cristina G. Carvalheiro, Suman Chaurasia, Viviane Rinaldi Favarin Colas, Simon Cousens, Ana Carolina Dantas de Assis, Han Dong, Angela Dramowski, Nguyen Trong Dung, Jinxing Feng, Youri Glupczynski, Srishti Goel, Herman Goossens, Doan Thi Huong Hao, Mahmudul Hasan, Tatiana Munera Huertas, Nathalie Khavessian, Angeliki Kontou, Tomislav Kostyanev, Premsak Laoyookhon, Sorasak Lochindarat, Maia De Luca, Surbhi Malhotra-Kumar, Nivedita Mondal, Nitu Mundhra, Philippa Musoke, Marisa M. Mussi-Pinhata, Ruchi Nanavati, Firdose L. Nakwa, Sushma Nangia, Alessandra Nardone, Borna Nyaoke, Christina W Obiero, Wang Ping, Kanchana Preedisripipat, Shamim Qazi, Lifeng Qi, Amy Riddell, Lorenza Romani, Praewpan Roysuwan, Robin Saggers, Samir Saha, Kosmas Sarafidis, Valerie Tusibira, Sithembiso Velaphi, Tuba Vilken, Xiaojiao Wang, Yajuan Wang, Yonghong Yang, Sally Ellis, Julia Bielicki, A Sarah Walker, Paul T. Heath, Mike Sharland

## Abstract

**Background:** Sepsis severity scores are used in clinical practice and trials to define risk groups. There are limited data to derive hospital-based sepsis severity scores for neonates and young infants in high-burden low- and middle-income country (LMIC) settings where trials are urgently required. We aimed to create linked sepsis severity and recovery scores applicable to hospitalized neonates and young infants in LMIC which could be used to inform antibiotic trials.

**Methods & Findings:** A prospective observational cohort study was conducted across 19 hospitals in 11 countries in sub-Saharan Africa, Asia, Latin America and Europe. Infants aged <60 days with clinical sepsis fulfilling at least two clinical or laboratory criteria (≥1 clinical) were enrolled. Primary outcome was 28-day mortality. Two prediction models were developed for 1) 28-day mortality from factors at sepsis presentation (baseline NeoSep Severity Score), and 2) daily risk of death on IV antibiotics from daily updated assessments (NeoSep Recovery Score). Multivariable Cox regression models included a randomly selected 85% of infants, with 15% for validation.

3204 infants were enrolled between 2018-2020. Median age was 5 days (IQR 2-15), 90.4% (n=2,895) were <28 days. Median birth weight was 2500g (1400-3000g), and a median of 4 clinical (IQR 2-5) and 1 laboratory (0-2) signs were present. Overall mortality was 11.3% (95%CI 10.2-12.5%; n=350). A baseline NeoSep Severity Score from infants characteristics, respiratory support, and clinical signs (no laboratory tests) at presentation had a C-index 0.77 (95%CI: 0.75-0.80) and 0.76 (0.69-0.82) in derivation and validation samples, respectively. Mortality in the validation sample was 1.6% (3/189; 95%CI: 0.5-4.6%), 11.0% (27/245; 7.7-15.6%), and 27.3% (12/44; 16.3-41.8%) in low (score 0-4), medium (5-8) and high (9-16) risk groups, respectively, with similar performance across subgroups.

A related NeoSep Recovery Score based on evolving post-baseline clinical signs and supportive care discriminated well between infants who died or survived the following day or subsequent few days. The area under the ROC curve for score on day 2 and death in the following 5 days was 0.82 (95%CI 0.78-0.85) and 0.85 (95%CI 0.78-0.93) in the derivation and validation data, respectively.

**Conclusion:** The baseline NeoSep Severity Score predicted 28-day mortality and could identify infants with high risk of mortality for inclusion in hospital-based sepsis trials. The NeoSep Recovery Score predicts day-by-day inpatient mortality and could, with further validation, help to identify poor response to antibiotics.

**Author Summary:** *Why was this study done?:*

➣ Evidence to guide hospital-based antibiotic treatment of sepsis in neonates and young infants is scarce, and clinical trials are particularly urgent in low- and middle-income (LMIC) settings where antimicrobial resistance threatens to undermine existing guidelines
➣ There is limited data to inform the design of antibiotic trials in LMIC settings, particularly to define risk stratification and inclusion and escalation criteria in hospitalised neonates and young infants

*What did the researchers do and find?:*

➣ To our knowledge this is the first global, prospective, hospital-based observational study of clinically diagnosed neonatal sepsis across 4 continents including LMIC settings, with extensive daily data collection on clinical status, antibiotic use and outcomes.
➣ There was a high mortality among infants with sepsis in LMIC hospital settings. 4 non-modifiable and 6 modifiable factors predicted mortality and were included in a NeoSep Severity score which defines patterns of mortality risk at baseline
➣ A NeoSep Recovery Score including the same modifiable factors (with the addition of cyanosis) predicted mortality on the following day during the course of treatment.

*What do these findings mean?:*

➣ The NeoSep Severity Score and NeoSep Recovery score are now informing inclusion and escalation criteria in the NeoSep1 antibiotic trial (ISRCTN48721236) which aims to identify novel first- and second-line empiric antibiotic regimens for neonatal sepsis
➣ The NeoSep Severity Score could be used to predict mortality at baseline in future studies of targeting resources in routine care. With further validation, the NeoSep Recovery Score could potentially be used to identify poor response to empiric antibiotic treatment

## Introduction

There are an estimated 1.3 million episodes of neonatal sepsis^1^ and 200,000 sepsis-attributable deaths each year, largely in low- and middle-income countries (LMIC).^2^ Increasing antimicrobial resistance (AMR), particularly in hospitals, threatens to undermine the effectiveness of available antibiotic treatment, with bacterial isolates increasingly resistant to World Health Organization (WHO) recommended first- and second-line regimens.^3–8^

In this context, global routine antibiotic use for neonatal sepsis frequently diverges from WHO guidelines.^9^ There are limited clinical data or scoring systems applicable to LMIC hospitals to stratify risk and predict mortality, assess response to treatment and guide decisions on antibiotic escalation and de-escalation.^10^ Most recent large-scale sepsis trials in neonates and young infants in LMIC have been community-based, focusing on simplifying antibiotic regimens,^11–13^ with very limited evidence available to optimize treatment in higher risk hospital settings where treatment and recovery is more complex.

Challenges to conducting antibiotic trials in neonates and young infants include lack of regulatory guidance and no widely accepted case definition for neonatal sepsis.^14^ European Medicines Agency (EMA) criteria for neonatal sepsis are comprehensive but perform variably^15^ and require laboratory tests which are not always available in LMIC. Criteria for possible serious bacterial infection (pSBI) are well adapted for LMIC but focus on community-acquired sepsis at primary care level and identify large numbers of infants with mild illness.^11–13^ There is a need for criteria to stratify risk and define inclusion criteria for trials in hospital-based populations of neonates and young infants.

The aim of this analysis was to develop two linked clinically based scores relevant to LMIC populations: 1) a sepsis severity score to predict 28-day mortality from factors known at sepsis presentation and 2) a recovery score to predict the daily risk of death on treatment with IV antibiotics using daily updated assessments of clinical status. Additional results of the NeoOBS cohort are published separately.

## Methods

### Study design and participants

Hospitalized infants aged <60 days with a new episode of clinically suspected sepsis were enrolled from 19 hospitals across 11 countries, in Asia (Bangladesh, China, India, Thailand, Vietnam), Africa (Kenya, South Africa, Uganda), Europe (Italy, Greece) and South America (Brazil). Sites were selected after a feasibility study^16^ to represent diverse regions and to include secondary and tertiary referral hospitals, public facilities, and facilities with varying proportions of in-born and out-born neonates. Access to blood cultures and high-quality microbiology was required.

Infants were eligible if the local physician had decided to treat the infant with antibiotics for a new distinct sepsis episode meeting the inclusion criteria (supplement figure 1), which combined clinical and laboratory criteria from WHO pSBI^17^ and EMA Criteria.^15^ To allow for variation in access to laboratory testing, and ensure generalizability to varying LMIC hospital contexts, laboratory values could be used for inclusion but were <u>not</u> mandatory. A minimum of 2 clinical, or 1 clinical and 1 laboratory, sepsis criteria were required with a locally adapted sampling frame used to recruit up to 200 infants per site. Infants were excluded if an alternative primary diagnosis other than sepsis was suspected, or a serious non-infective comorbidity was expected to cause death within 72 hours.

Ethical approval was obtained from St. George’s, University of London (SGUL) Research Ethics Committee and sites’ local, central or national ethics committees and other relevant local bodies, where required. Design and reporting were guided by the STROBE-NI framework.^18^

### Procedures

A blood culture taken before new antibiotics were started was the only compulsory procedure. After written informed consent from parents/guardians, with a witness for those who could not read/write, data were collected on demographics, maternal history, birth history, pre-existing comorbidities and previous infections, as well as available clinical data from 24 hours before the time of blood culture sampling. Infants were prospectively followed with daily observations recorded on vital signs, clinical signs and supportive treatment.

Infants were followed for 28 days after enrolment in-person if still hospitalized, or by telephone post-discharge. A final diagnosis was documented by clinicians, as was primary and secondary causes of death, and any clinical illness or readmission occurring after discharge and within 28 days of enrolment. ‘Infection-related death’ was independently classified by the research team based on isolation of a presumed pathogen from blood or CSF culture, reporting by clinicians of infection as a cause of death, or a mode of death consistent with infection in the absence of another reported cause.

Data were collected by research and clinical staff based on clinical observation and routine source documentation (e.g. medical/nursing notes, vital signs and prescription charts), and entered and managed using REDCap(tm) electronic data capture tools^19^ hosted at SGUL.

### Statistical analysis

The pre-specified primary outcome was mortality through 28 days post-enrolment. We developed two prediction models and corresponding risk scores: 1) a baseline NeoSep Severity Score to predict 28-day mortality from factors known at sepsis presentation, and 2) a NeoSep Recovery Score to predict the daily risk of death while treated with IV antibiotics from daily updated assessments of clinical status. For both analyses, a randomly selected 15% of infants per site was reserved for model validation and not used in model development. In the remaining 85% of infants, Cox proportional hazards regression with site-level random effects were used, with time measured from the initial blood culture sample, censoring at the earliest of day 28, withdrawal or last contact if lost post-discharge, or, for the recovery analysis, when IV antibiotics stopped (cause-specific model). Analyses used Stata version 16.1.

#### NeoSep Severity Score

Backwards elimination (exit p=0.05) was used to identify independent predictors of 28-day mortality from a pre-defined set of factors including infants’ characteristics, supportive care and clinical signs that could be considered known at sepsis presentation (Table 1). A points-based risk score integerised model coefficients for selected predictors, with higher scores indicating higher mortality risk. Discrimination was assessed using Harrell’s C-index with bootstrapped confidence intervals. The NeoSep Severity Score was compared with a score based on WHO pSBI^17^ allocating one point for every criterion.

**Table 1:**
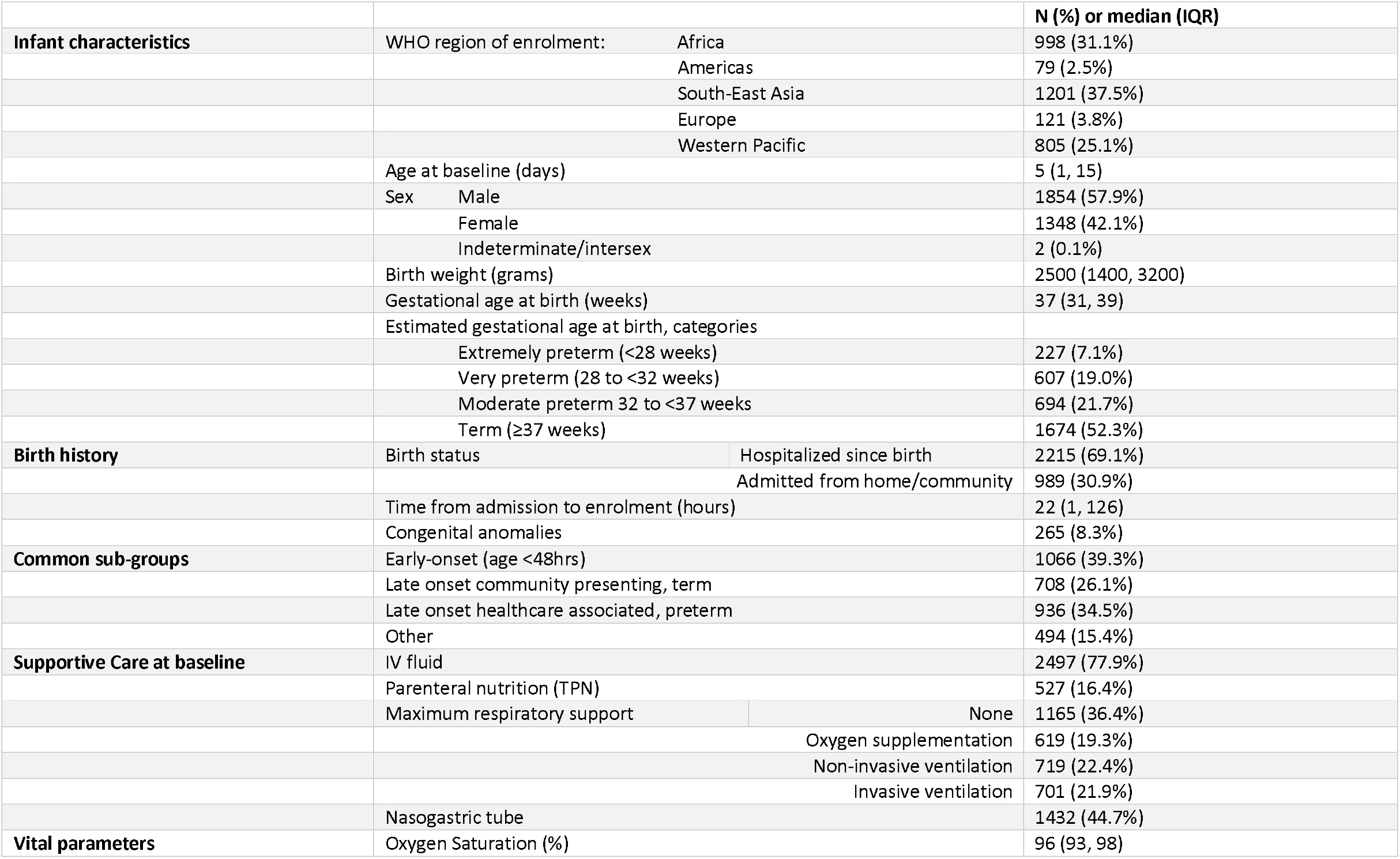
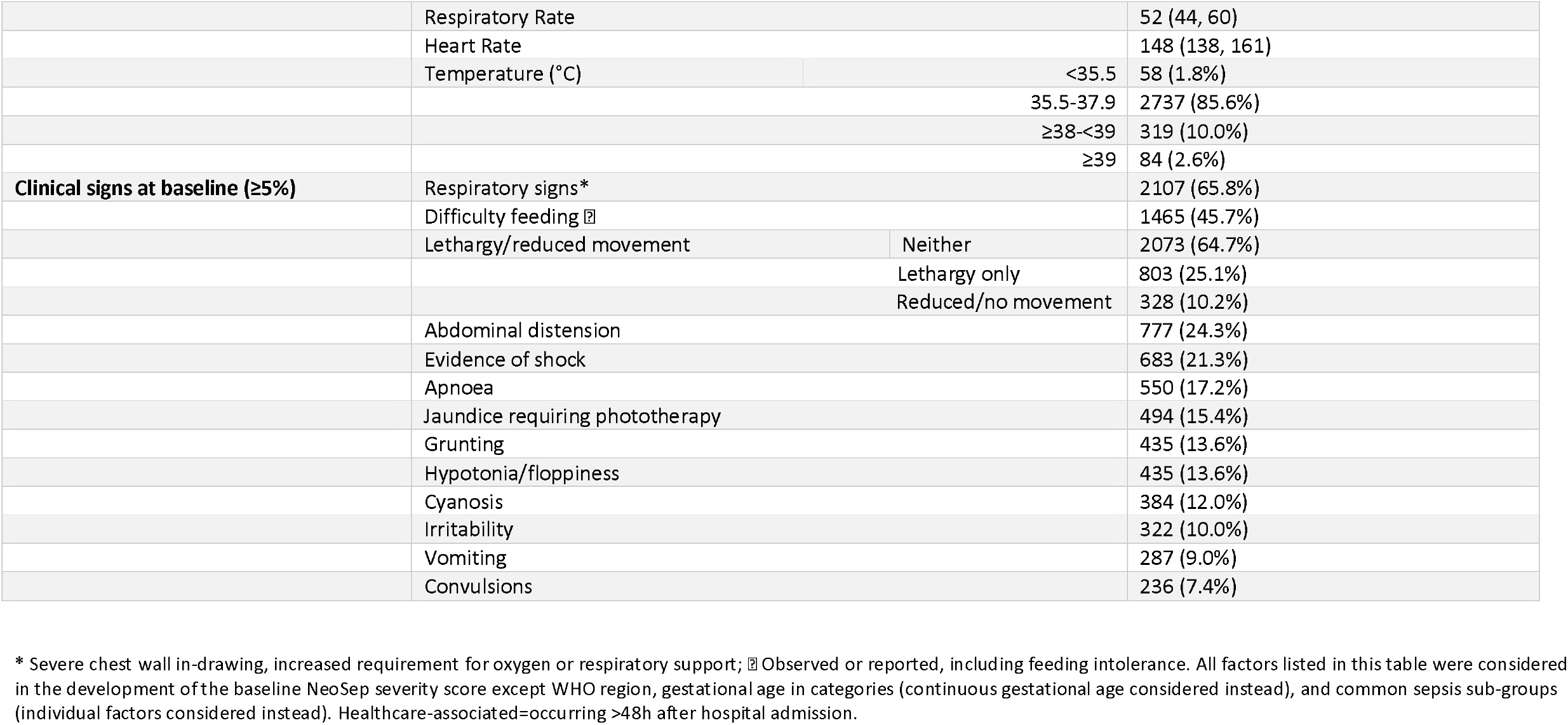
Baseline characteristics.

#### NeoSep Recovery Score

To estimate the daily risk of death after initiating IV antibiotics, we initially included all clinical factors in the baseline NeoSep severity score as time-updated factors, regardless of significance; excluding unmodifiable infant and birth characteristics, such as gestational age, that could not evolve and therefore could not reflect recovery. Forward selection (entry p=0.05) was then used to identify additional independent time-updated clinical predictors. To avoid selecting factors representing the mechanism of dying rather than being predictors of subsequently dying, clinical parameters reported on a given day were used to predict death on the following day in all time-updated models. A points-based risk score was then derived similarly to the baseline severity score. Discrimination was assessed using time updated area under the receiver operating curve (AUROCs). To examine the potential usefulness of the recovery score for informing the decision to potentially switch to second line antibiotics in future empiric clinical trials, we focused on day 2 (48-72 hours) post baseline, a key decisional time point during clinical management when culture results become available and response to treatment is commonly evaluated (see supplement statistical methods for details).

## Results

3204 infants (90.4% neonates aged <28 days, n=2895; 42.1% female, n=1348) were recruited from 20^th^ August 2018-29^th^ February 2020. The median postnatal age was 5 days (IQR 1-15). 3088 (96.4%) infants had been born in a hospital/facility (1550 in the enrolling facility), 1412 (44.3%) by caesarean section (969 as an emergency). The median (IQR) gestational age at birth was 37 (31-39) weeks, with birth weight 2500g (1400-3000g). When enrolled, 69.1% (n=2215) infants had been hospitalized since birth, and 30.9% (n=989) were admitted from the community. 2759 (86.1%) were recruited in a neonatal unit (supplement table 1). 41.1% (n=1318) of sepsis episodes were healthcare-associated (occurring >48h after hospital admission). Among 309 (9.6%) infants enrolled aged ≥28 days, the majority (n=181; 58.6%) were ex-premature (n=146; 47.4%) and/or had been admitted since the neonatal period (n=136, 44.2%), the majority of which since birth (n=136, 86.8%). Other than infection, the most common diagnoses/problems reported at admission were respiratory distress (51.4%), prematurity (41.1%) and low birth weight (36.5% (supplement figure 2)). Supportive measures such as invasive ventilation varied across sites, as described in supplement figure 3.

A median of 4 (IQR 2-5) clinical signs were present at baseline, the most common being respiratory signs (65.8%, n=2107), difficulty feeding (45.7%, n=1464), lethargy or reduced movement (35.3%, n=1,131), abdominal distension (24.3%, n=777) and evidence of shock (21.3%, n=683), with prevalence decreasing over time (table 1 & supplement figure 4). Signs associated with meningitis were relatively uncommon (irritability 10% (n=322), convulsions 7.4% (n=236), abnormal posturing 4.4% (n=140), bulging fontanelle 1.8% (n=57)) (supplement figure 5). Initial blood culture results were available a median of 2 days (IQR 1-3) after blood draw for 3195 (99.7%) infants, among whom pathogens were isolated in 564 (17.6%) (>1 pathogen in 29).

### Mortality

Overall, 350 infants (11.3%; 95%CI 10.2-12.5%) died within 28 days of the baseline blood culture, with wide variation between sites (1.0% to 27.6%, supplement figure 6). Mortality among infants with a pathogen-positive baseline culture was 17.7% (99/564; 95%CI 14.7-21.1%) compared with 9.9% (250/2631; 8.8-11.2%) in infants without pathogens. 315/350 infants died in the initial hospital (286 whilst still on IV antibiotics), and 35 after discharge (35/350, 10.0%). Reported causes of death included infection in 74.9% (n=262), prematurity in 48.0% (n=168), birth asphyxia in 10.6% (n=37) and congenital malformations in 10.3% (n=36), with infection and prematurity being the most common concomitant diagnoses. After standardized review, 309 (88.3%) deaths were classified as ‘infection-related’. Mortality at 28 days was unknown in 62 infants (5 withdrew in hospital, 57 were lost post-discharge).

### Baseline predictors of 28-day mortality

Ten clinical factors known at presentation independently predicted mortality in the final model, including infant characteristics (birth weight, gestational age, duration of time in hospital, congenital anomalies), level of respiratory support, and clinical signs (abnormal temperature, abdominal distension, lethargy/no or reduced movement, difficulty feeding, and evidence of shock) (table 2). Risk was increased with both low (<35°C) and high temperature (≥38°C) with evidence of even higher risk if ≥39°C. A NeoSep Severity Score developed from these baseline predictors had a maximum 16 points (table 2), with C-statistics 0.77 (95%CI: 0.75-0.80) and 0.76 (0.69-0.82) in the derivation and validation samples, respectively, and a good fit in the validation sample (Hosmer-Lemeshow p=0.53). Defining mortality risk thresholds at 5% and 25% in the derivation sample categorized scores as low (score 0-4), medium (5-8) and high (9-16) risk, with mortality in the derivation sample of 1.6% (17/1071; 95%CI: 1.0-2.5%), 14.4% (203/1409; 12.7-16.3%), and 35.8% (88/246; 30.0-41.9%), and in the validation sample of 1.6% (3/189; 95%CI: 0.5-4.6%), 11.0% (27/245; 7.7-15.6%), and 27.3% (12/44; 16.3-41.8%)(figure 1).

**Table 2:** Predictors of mortality and risk score.

| Factor |  | NeoSep Severity Score (at presentation) |  |  |  | NeoSep Recovery Score (daily on IV antibiotics) |  |  |  |
| --- | --- | --- | --- | --- | --- | --- | --- | --- | --- |
|  |  | HR (95% CI) | p-value | Coef | Severity score points<sup>†</sup> | HR (95% CI) | p-value | Coef | Recovery score points<sup>†</sup> |
| Birth weight (kg) <sup>‡</sup> | 1.0 | 2.13 (1.55,2.92) | <0.001 | 0.67 | <1 kg: 2 |  |  |  |  |
|  | 2.0 | 1.21 (1.12,1.31) |  | 0.25 | 1-2 kg: 1 |  |  |  |  |
|  | 3.0 | ref |  | 0 |  |  |  |  |  |
| Time in hospital | per additional 24 hours | 0.96 (0.94,0.98) | <0.001 | -0.04 | ≤ 10 days: 1 |  |  |  |  |
| Gestational age | per additional week | 0.94 (0.90,0.98) | 0.004 | -0.06 | < 37 weeks: 1 |  |  |  |  |
| Congenital anomalies |  | 2.81 (1.93,4.08) | <0.001 | 1.03 | 2 |  |  |  |  |
| Maximum respiratory support: | Oxygen supplementation | 2.87 (1.74,4.74) | <0.001 | 1.06 | 2 | 9.21 (4.04,21.0) | <0.001 | 2.22 | 2 |
|  | CPAP, BiPAP, HFNC | 4.72 (2.78,8.00) | <0.001 | 1.55 | 3 | 15.2 (6.65,34.8) | <0.001 | 2.72 | 3 |
|  | Invasive ventilation | 10.2 (6.05,17.1) | <0.001 | 2.32 | 3 | 48.6 (21.5,110) | <0.001 | 3.88 | 3 |
| Temperature (°C) <sup>‡</sup> | 35.5 | 1.17 (0.78,1.75) | <0.001 | 0.15 | <35.5 °C: 1 | 1.32 (0.97,1.79) | 0.005 | 0.26 | <35.5 °C: 1 |
|  | 37 | ref |  | 0 |  | ref |  | 0 |  |
|  | 38 | 1.41 (1.18,1.69) |  | 0.35 | ≥38 – <39 °C: 1 | 1.43 (1.07,1.92) |  | 0.34 | ≥38 – <39 °C: 1 |
|  | 39 | 2.91 (1.76,4.81) |  | 1.07 | ≥39°C: 2 | 3.07 (1.50,6.31) |  | 1.11 | ≥39°C: 2 |
| Abdominal distension |  | 1.60 (1.24,2.07) | <0.001 | 0.47 | 1 | 1.50 (1.13,2.00) | 0.005 | 0.41 | 1 |
| Lethargy, no or reduced movement: | Lethargy only | 1.28 (0.97,1.69) | 0.083 | 0.25 | 1 | 1.85 (1.33,2.59) | <0.001 | 0.62 | 1 |
|  | No/reduced movement (± lethargy) | 2.00 (1.36,2.95) | <0.001 | 0.70 | 2 | 3.10 (2.08,4.61) | <0.001 | 1.13 | 2 |
| Difficulty feeding |  | 1.50 (1.14,1.98) | 0.004 | 0.41 | 1 | 2.15 (1.56,2.95) | <0.001 | 0.76 | 1 |
| Evidence of shock |  | 1.69 (1.28,2.24) | <0.001 | 0.53 | 1 | 2.51 (1.81,3.48) | <0.001 | 0.92 | 1 |
| Cyanosis |  |  |  |  |  | 1.87 (1.25,2.78) | 0.002 | 0.62 | 1 |
| Maximum number of score points possible |  |  |  |  | 16 |  |  |  | 11 |

Notes: <sup>‡</sup> Birth weight and temperature were analysed as continuous variables using fractional polynomials with powers -2 for birth weight, and powers 2 2 for temperature. For illustrative reasons, in this table we report HRs for specific values.<sup>†</sup> For each model, score points for each factor were derived by dividing its coefficient by the smallest significant coefficient of any categorical factor, rounding to the nearest integer, and giving 1 score point for <3 raw points, 2 score points for 3-5 raw points, and 3 score points for ≥6 raw points. Shaded factors: not a significant predictor (cyanosis: baseline severity score) or deliberately not included in model (unmodifiable factors excluded from recovery score: birth weight, time in hospital, gestational age, congenital anomalies). CPAP = Continuous Positive Airway Pressure, BiPAP Bilevel Positive Airway Pressure, HFNC=High Flow Nasal Cannula.

**Figure 1:**
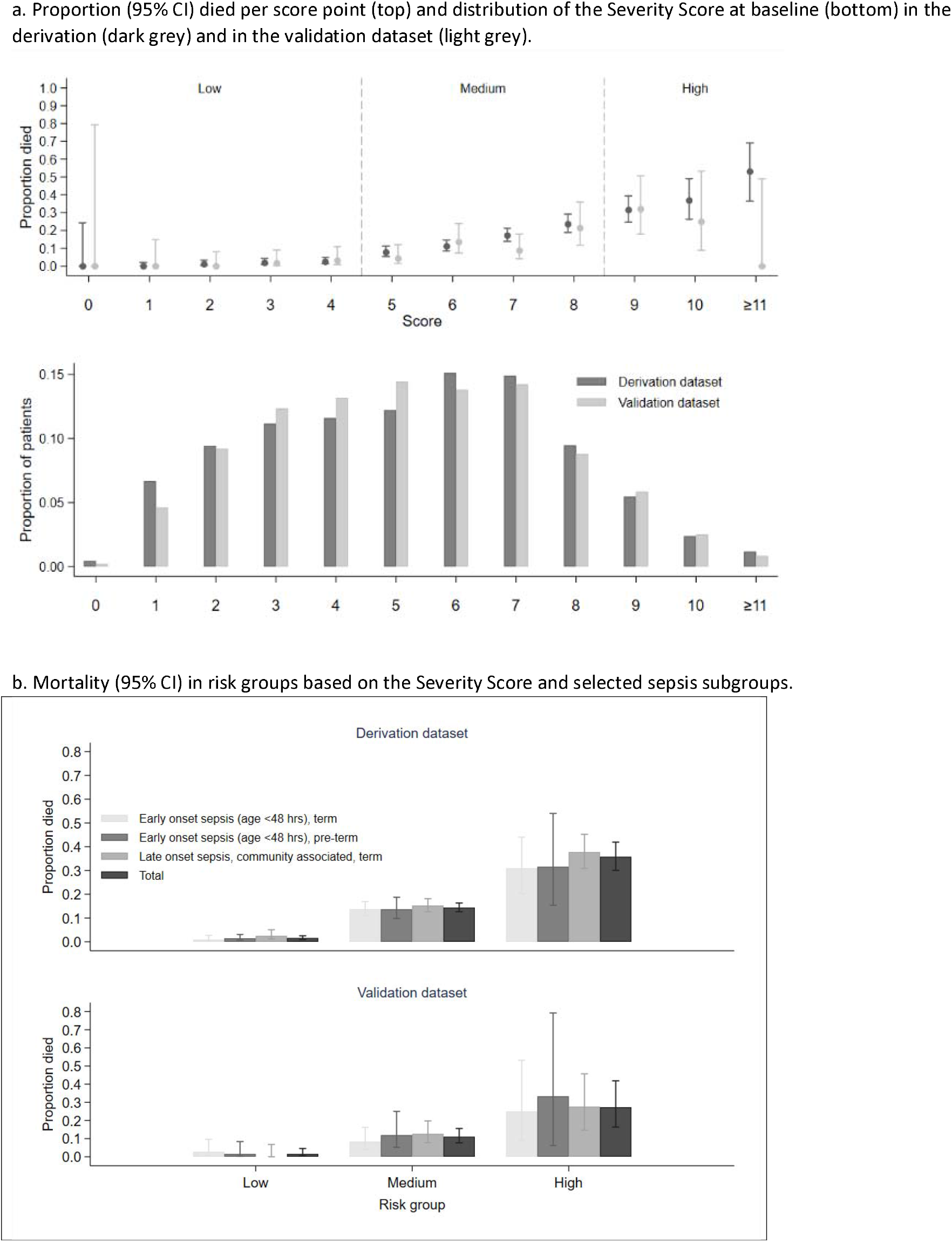
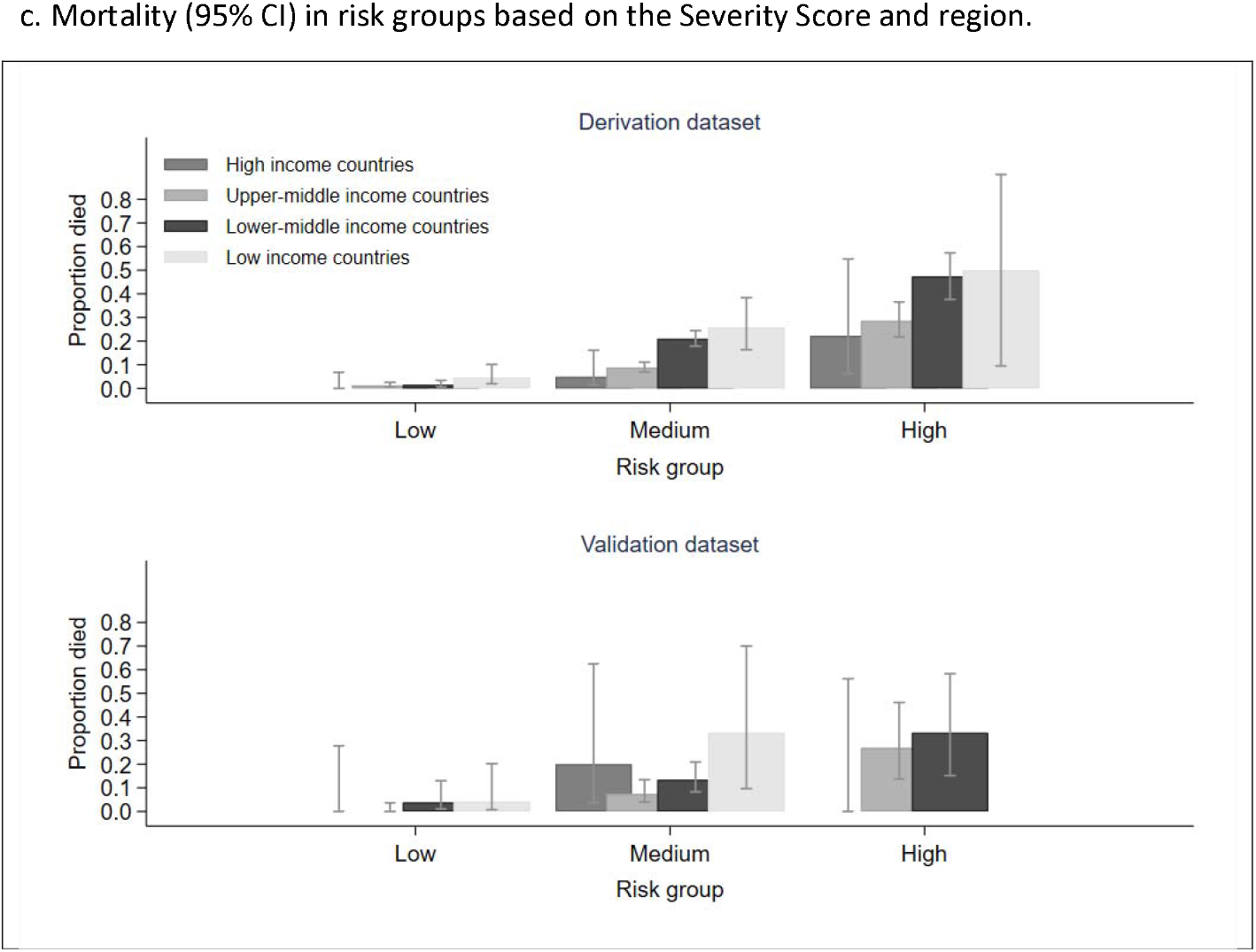
NeoSep Severity Score at baseline.

The association between the NeoSep Severity Score and mortality was similar within multiple sub-groups, for example in early or late-onset, community or healthcare-associated, high-, middle- or low-income settings (figure 1), term vs. preterm infants, blood culture-positive vs. culture-negative cases (supplement figure 7).

A score based on WHO pSBI had a C-statistic of 0.63 (95%CI 0.56-0.70) in the validation sample, lower than the NeoSep Severity Score. Because the WHO pSBI do not include any infant/birth characteristics, to ensure an appropriate comparison we also calculated a modified NeoSep Severity Score excluding these factors which had a C-statistic of 0.72 (95%CI 0.65-0.77) (supplement figure 8).

### Predictors of daily risk of dying of sepsis while on intravenous antibiotics in hospital: NeoSep Recovery Score

Seven of the time-varying factors independently predicted mortality in the final model, including all the non-modifiable signs in the NeoSep Severity Score (respiratory support and the five clinical signs), plus cyanosis, which had not added independent information in the baseline model (p=0.228) (table 2). A NeoSep Recovery Score developed from these time-updated predictors had a maximum 11 points and discriminated well between infants who died or survived the next day (figures 2&3). The area under the receiver operating curve (AUROC) of the recovery score on one day, for predicting death the next day, ranged between 0.8 and 0.9 over the first week post baseline in the derivation sample (figure 3). As expected, it slowly decreased over subsequent days. AUROC over time in the validation sample was similar although based on fewer numbers (figure 3).

**Figure 2:**
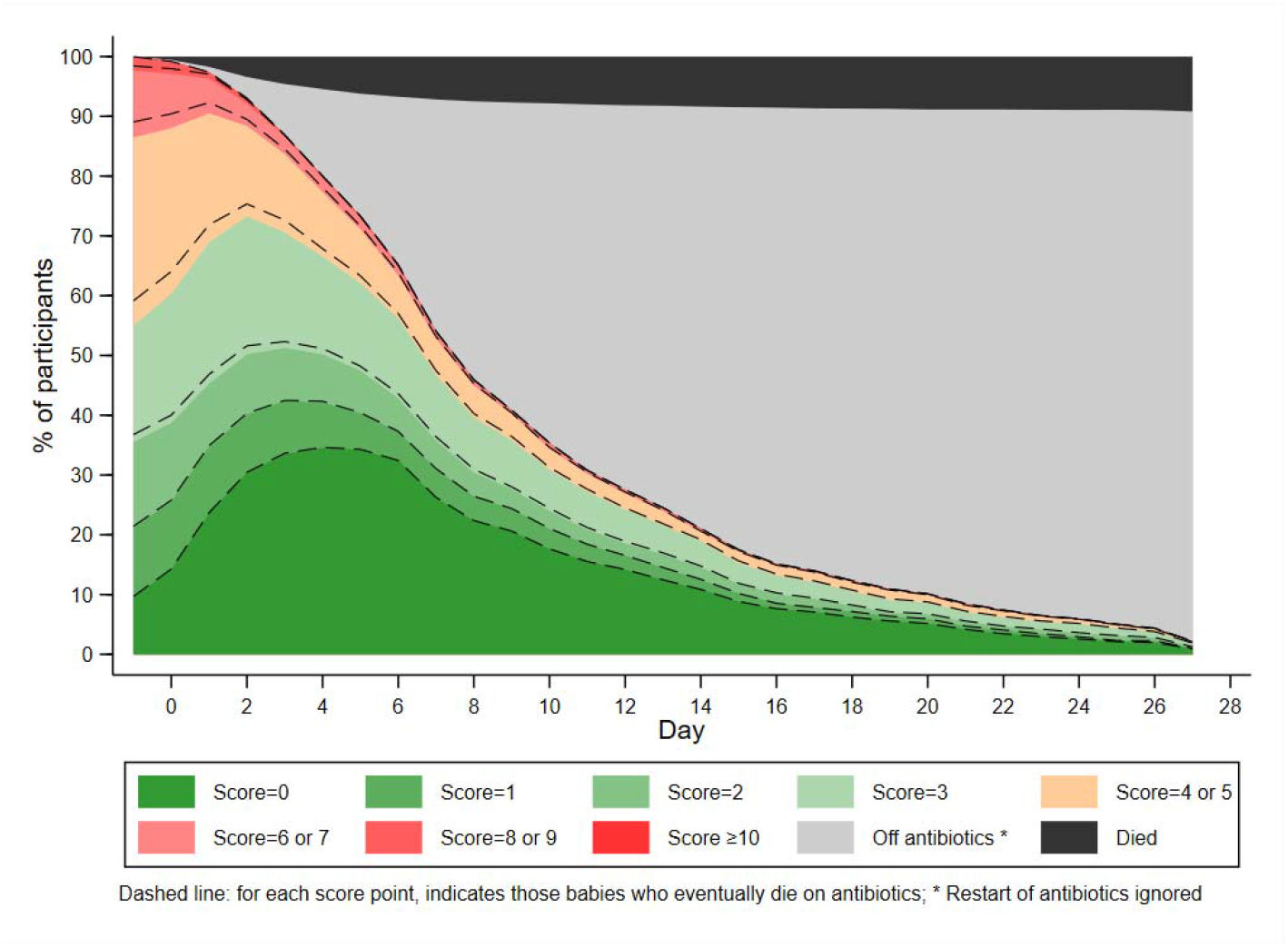
NeoSep Recovery Score over time (cross-sectional analysis).

**Figure 3:**
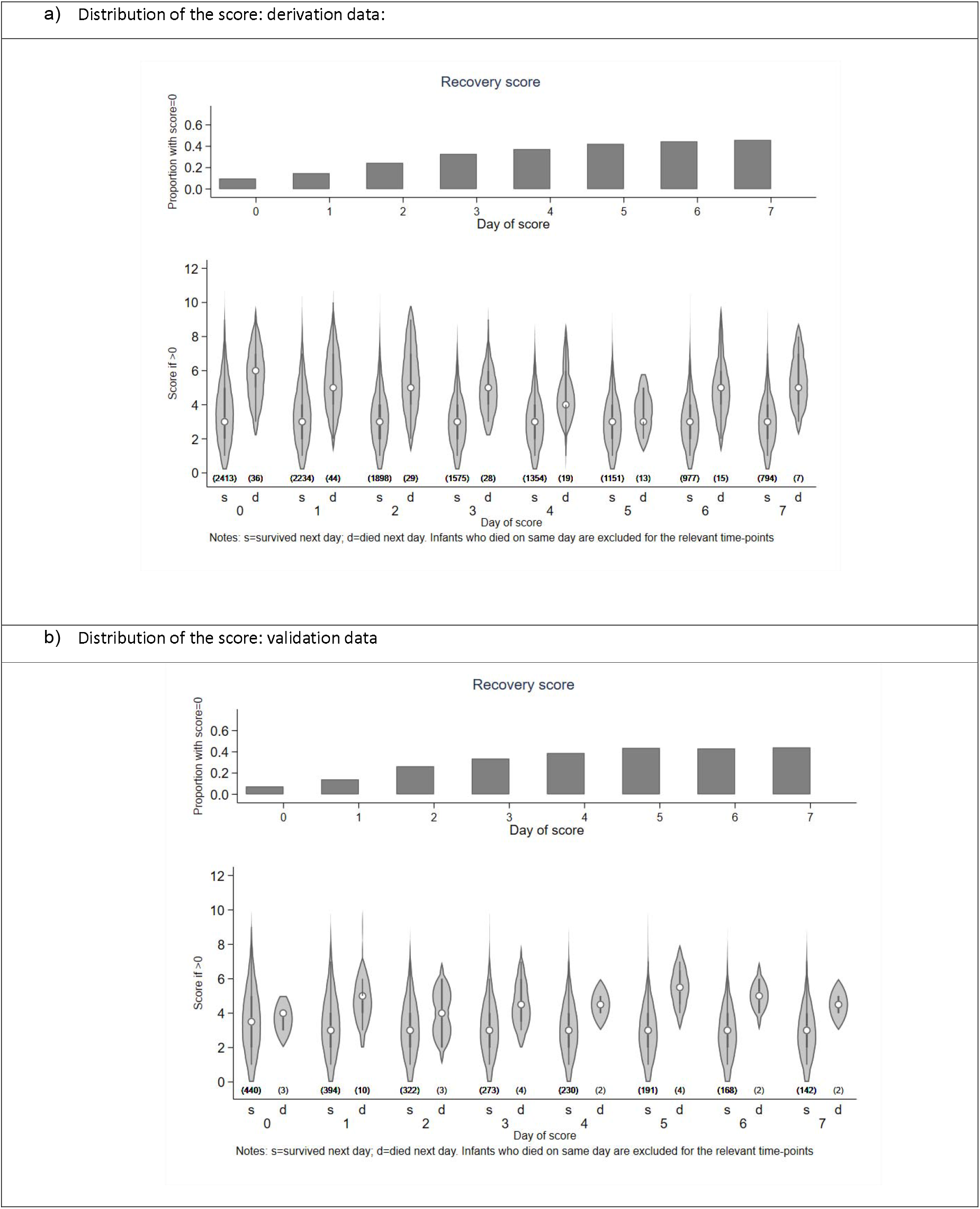
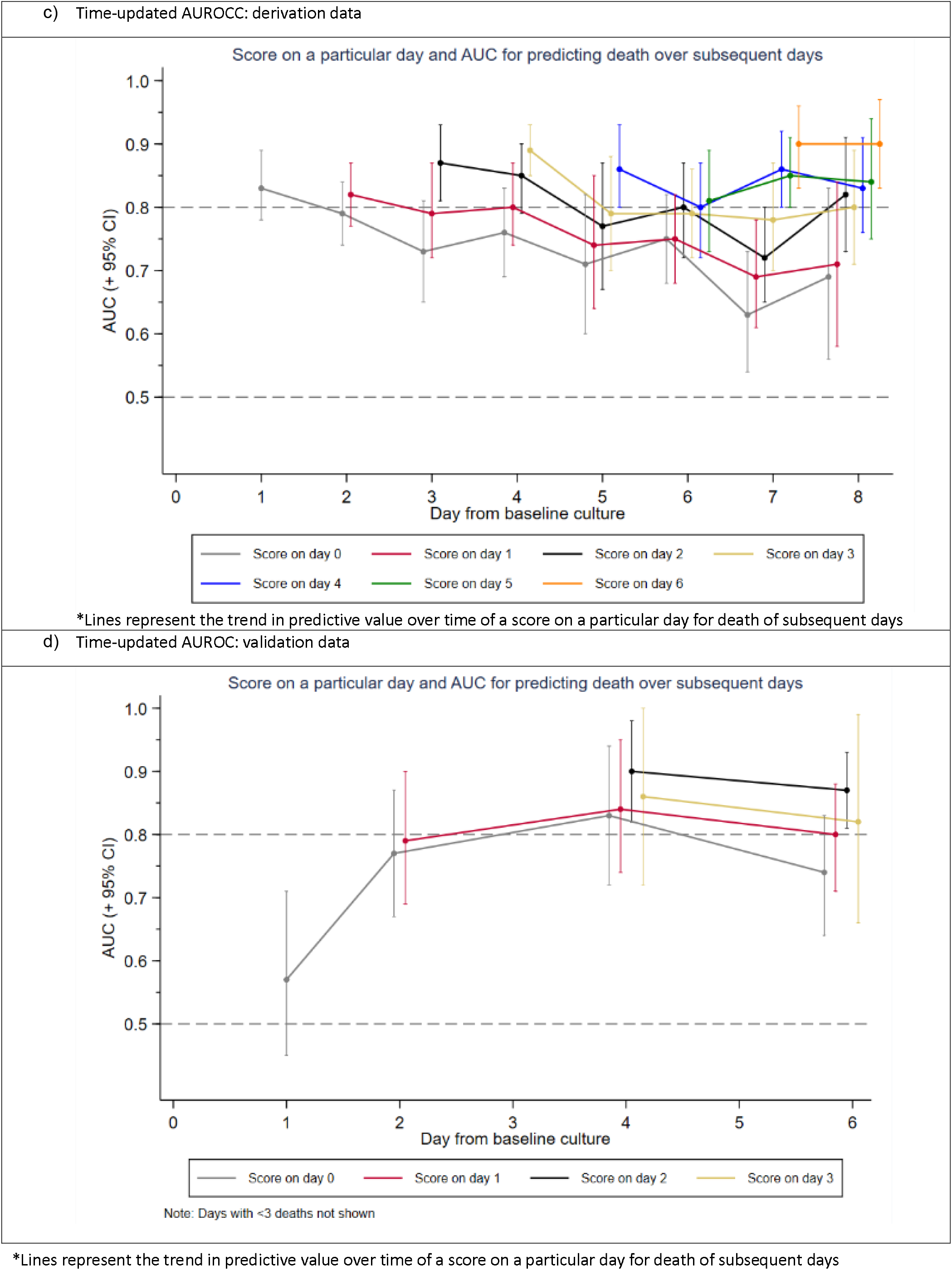
NeoSep Recovery Score.

The NeoSep Recovery Score on day 2 had an AUROC for dying in the following 5 days of 0.82 (95%CI 0.78-0.85) and 0.85 (95%CI 0.78-0.93) in the derivation and validation samples, respectively. A score ≥4 was the most discriminative, whether this was an increase from baseline, lack of initial response or an improvement from a higher score down to 4, with sensitivity and specificity of 0.74 (95%CI 0.64-0.82) and 0.74 (0.72-0.75) in the derivation, and 0.87 (0.60-0. 98) and 0.76 (0.71-0.79) in the validation samples.

Twenty-nine infants (derivation: n=27; validation: n=2) died between day 3 and 7 despite having had a score <4 on day 2. Of these, 6 deaths were not classified as infection-related, 10 had a subsequent increase in score to ≥4 before dying, 9 had at least 2 unmodifiable risk factors included as predictors of mortality in the NeoSep Severity Score (most commonly congenital abnormalities and very low birthweight), and 1 had congenital varicella; the remaining 4 had a day 2 score of 2 or more.

Of note, the change in score from baseline to day 2 had poorer discrimination than the absolute score on day 2, with an AUROC for dying in the following 5 days of 0.68 (95%CI 0.63-0.73) and 0.62 (95%CI 0.48-0.77) in the derivation and validation samples, respectively. Combining change in score using various cut-offs with <4/≥4 absolute score on day 2 also did not improve discrimination (supplement figure 9).

## Discussion

NeoOBS represents the largest hospital-based multi-country prospective observational cohort study to include high quality daily clinical data and outcomes in neonates and infants with sepsis in predominantly (>95%) LMIC settings. The cohort was largely neonatal (90%), with most of the remainder (>2/3) born prematurely and/or previously admitted during the neonatal period. All infants had a primary clinical diagnosis of sepsis and most deaths (88%) were infection-related. Overall mortality was 11.3%, with significant variation across different settings.

Mortality was independently associated with a range of factors, developed into a baseline sepsis severity score including unmodifiable infant characteristics and clinical characteristics at presentation/antibiotic initiation. A recovery score was developed accounting for evolving clinical signs after baseline. The scores use only simple clinical characteristics identifiable from routine history and examination and showed good discrimination. A score of 5 or higher at baseline was associated with 28-day mortality over 10%, and a recovery score 2 days after antibiotic initiation of 4 or higher had both sensitivity and specificity of 74% for mortality over the following 5 days. The baseline and recovery scores have informed the criteria for inclusion and escalation of therapy in the NeoSep1 antibiotic trial (ISRCTN48721236) for neonatal sepsis with a Personalised RAndomised Controlled Trial (PRACTical) design.^22^

To our knowledge, these are the first scores to specifically predict mortality risk developed from inpatient neonatal and young infant populations in multiple LMIC hospitals. While several single-centre studies in LMIC have identified individual risk factors for culture positivity and mortality in septic neonates,^23^ previous severity scores derived from hospital-based LMIC cohorts have been based on smaller studies assessing general illness severity rather than sepsis specifically.^24–29^ High-income setting based general illness severity scores are widely used,^24–26^ and some have been assessed in small populations of septic neonates.^30,31^ The nSOFA score is a recent sepsis-specific score developed on 60 very low birth weight (<1500g) infants, but relies on measurement of parameters which are often unavailable in LMIC settings (e.g. use of inotropes/vasoactive drug use).^32^

The NeoSep Severity and Recovery scores include clinical signs which have been designed to closely align with the WHO pSBI criteria (compared in supplement table 2) used in recent community-based studies, including AFRINEST^11^ and SATT.^12,13^ However, the NeoSep scores include additional factors more relevant to hospital settings which offer further predictive value, such as clinical evidence of shock, and need for oxygen and/or respiratory support. Importantly, the study population also included a more heterogenous mix of both preterm (including <1500g) and term infants, with early and late onset, and community and healthcare-associated sepsis, and the severity and recovery scores performed similarly across all these subgroups. Convulsions were uncommon in the study (7%) and were not associated with mortality despite being a pSBI criteria for critical illness, potentially due to low power.

The hospital sites included in this cohort were diverse, which is both a strength and potential limitation. Routine clinical practice was determined by sites, and there was variation in access to supportive care, with some sites using invasive ventilation and inotropic support and others with only oxygen and no access to vasoactive medication. Variation in intensity of clinical monitoring may also have influenced ascertainment of some factors such as shock, particularly as invasive monitoring was unavailable in some sites. Patient populations also varied between sites, as some were tertiary neonatal units, others treated more general populations, and there was variation in proportions of in-born versus out-born neonates. We attempted to account for inter-site variation by using multivariable models with site-level random-effects, excluding factors with a high proportion of missing values. We also examined performance of the severity score across different sites in high-, middle- and low-income contexts and demonstrated reproducible risk trends across all settings. Additional limitations of this study include the potential for selection bias. Prospective enrolment at the time of sepsis diagnosis may bias towards particular populations for whom parental consent is most practical, potentially leading to a milder phenotype than retrospective studies. Furthermore, the scores were validated on a predefined reserved 15% internal sample, including only 42 deaths (and relatively few infants with high-risk scores). External validation would strengthen the applicability of the scores. Nevertheless, inter-site variation reflected the diverse contexts across LMIC, and the data are likely to be representative of populations and subgroups that could feasibly be included in prospective antibiotic trials and subsequent global guideline development.

Overall mortality was high in the NeoOBS cohort of hospitalized neonates and young infants with sepsis in predominantly LMIC settings, with significant variation across different sites, highlighting the importance of standardised criteria to define severity. The NeoSep Severity and Recovery scores developed in this analysis define high-risk populations with clinical sepsis at treatment initiation and during recovery which can inform future research. In addition, the NeoOBS study demonstrates the feasibility of developing global hospital networks for conducting future trials focusing on improving outcomes through optimized prevention and treatment of sepsis in neonates and young infants.

## Data Availability

Requests for data should be adressed to GARDP

