## Supplementary appendix for "The NeoSep Severity and Recovery scores to predict mortality in hospitalized neonates and young infants with sepsis derived from the global NeoOBS observational cohort study"

[**5.**	**Supplement figure 5: Clinical signs at enrolment (<5%)**	4](#_Toc104677163)

1. **Supplement figure 1: Clinical and laboratory sepsis enrolment criteria**

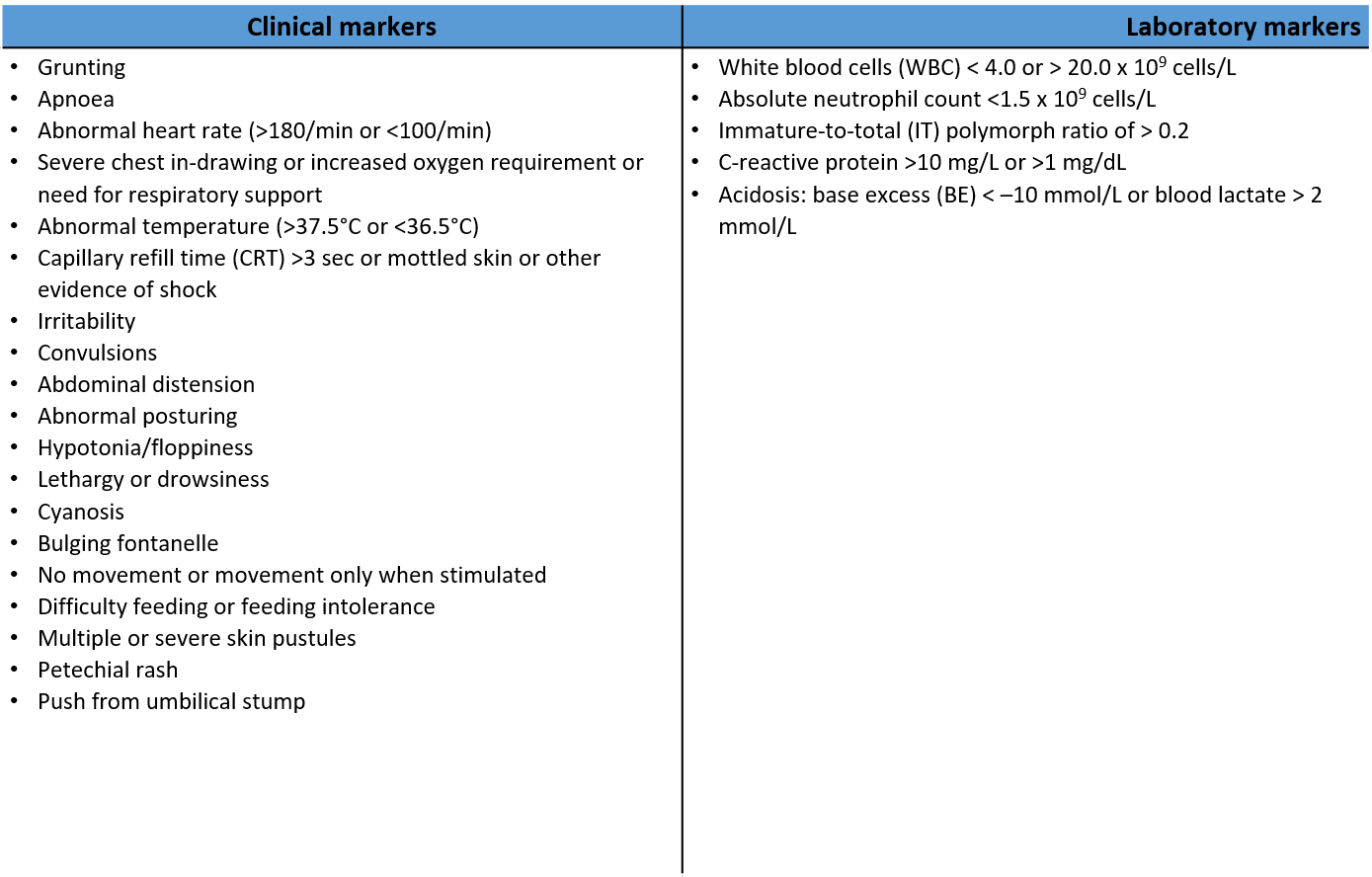

***** Primary suspected diagnosis should be significant sepsis - there should be no alternative explanation for these criteria (e.g. hypoxic ischaemic encephalopathy)

****** Infants must meet **two** criteria, **one** of which must be clinical

2. **Supplement figure 2: Diagnoses at time of Hospital Admission**

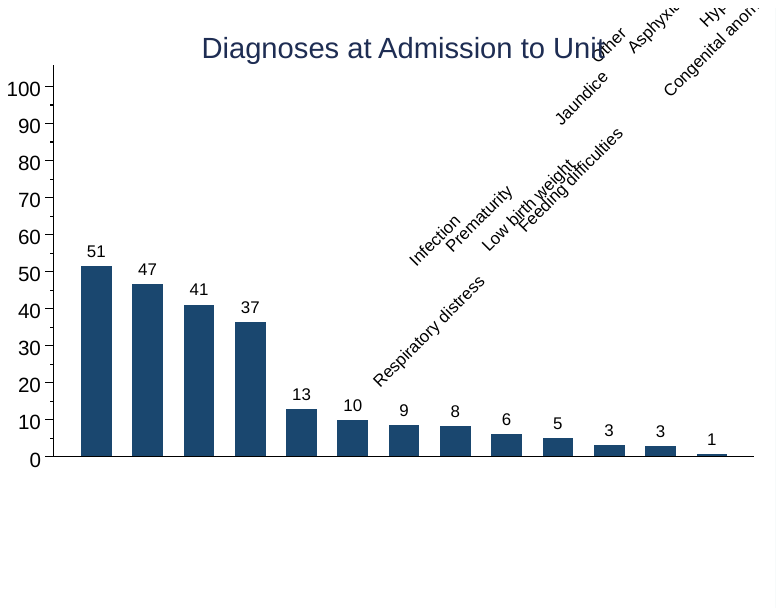

3. **Supplement figure 3: Frequency of supportive measures across sites**

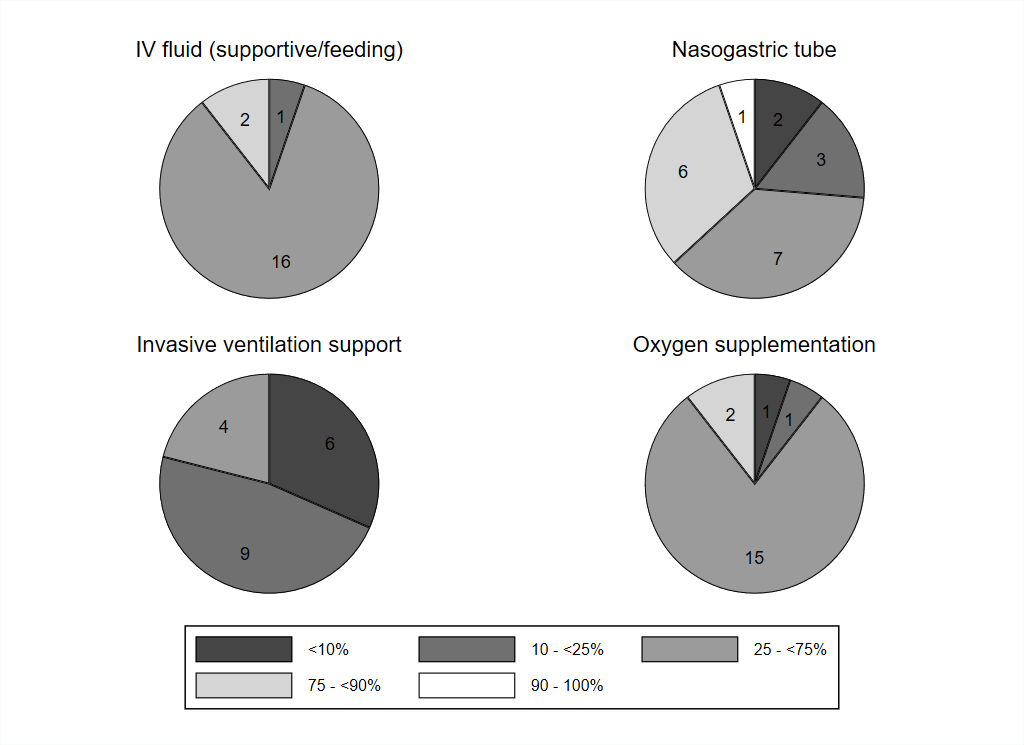

Note: Pictured are the number of sites with supportive measures performed in % of infants enrolled. Overall number of sites: n=19.

4. **Supplement figure 4: Prevalence of clinical signs / respiratory support over time**

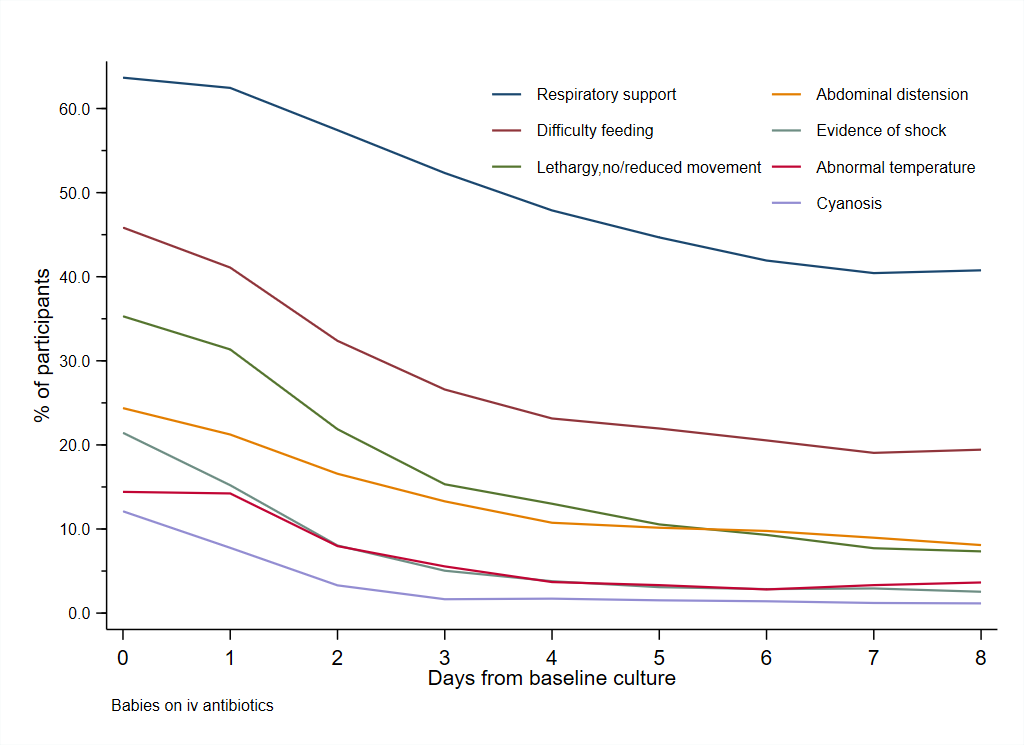

5. **Supplement figure 5: Clinical signs at enrolment (<5%)**

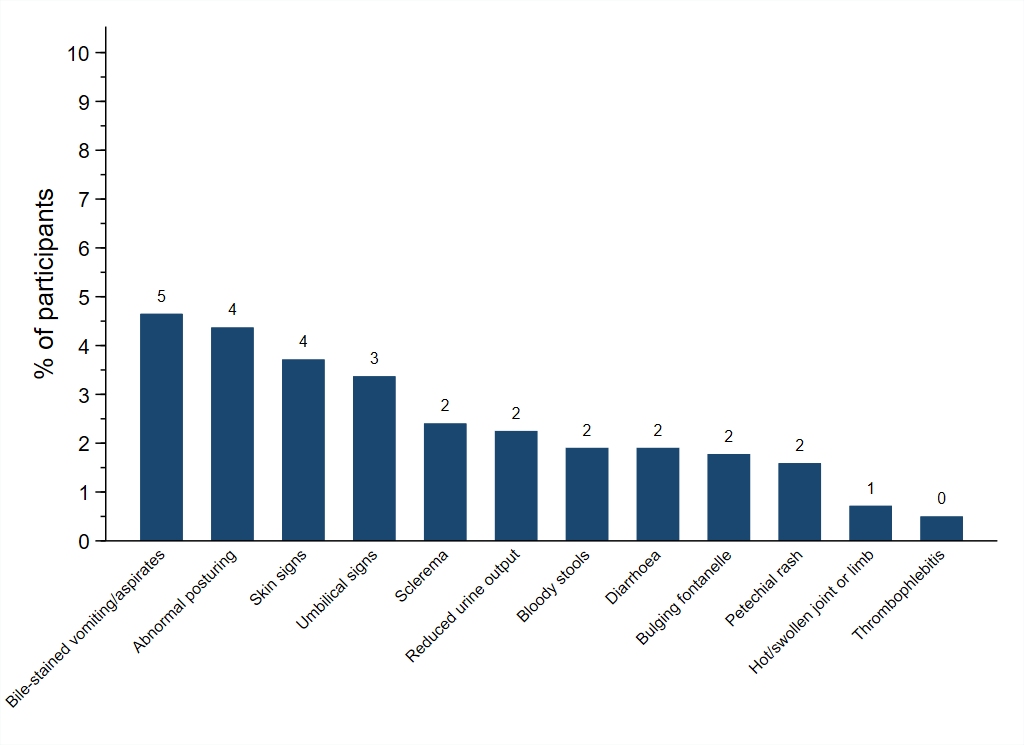

6. **Supplement figure 6: Kaplan-Meier curves for mortality, overall and by site**

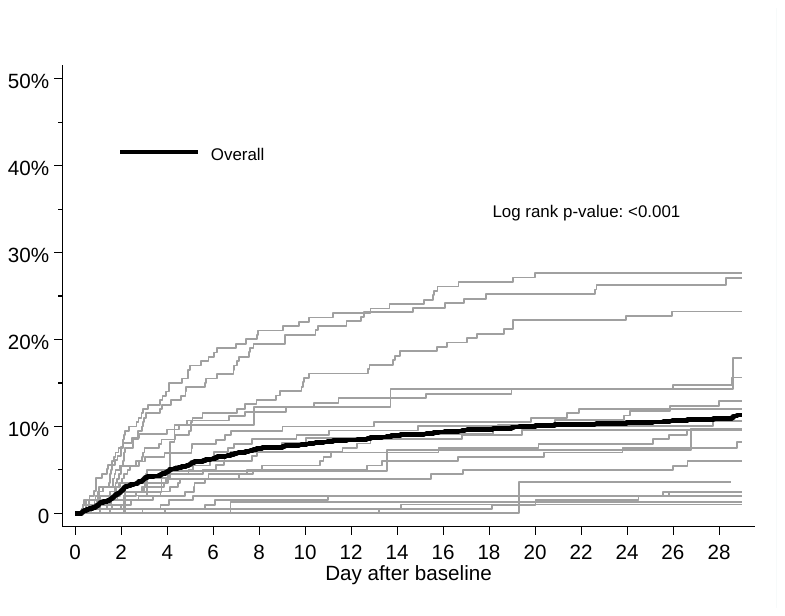

7. **Supplement figure 7: NeoSep Severity Score performance in subgroups**

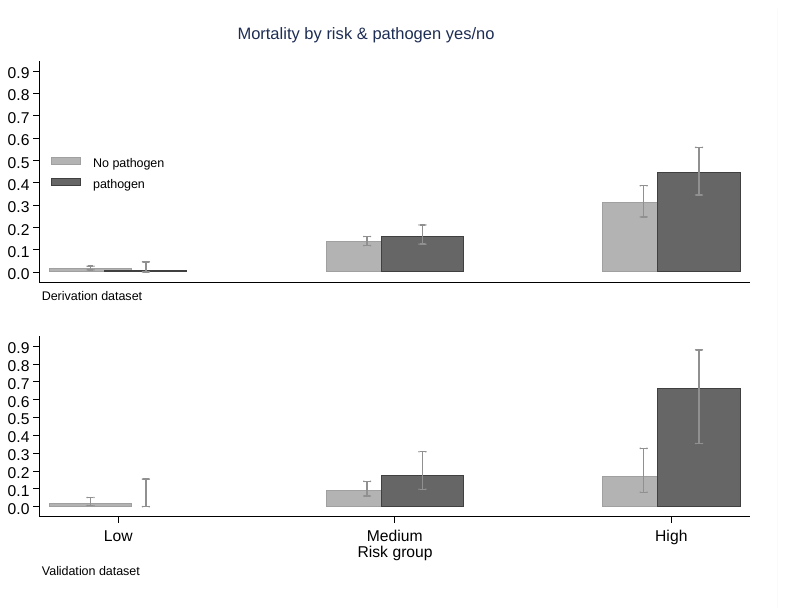

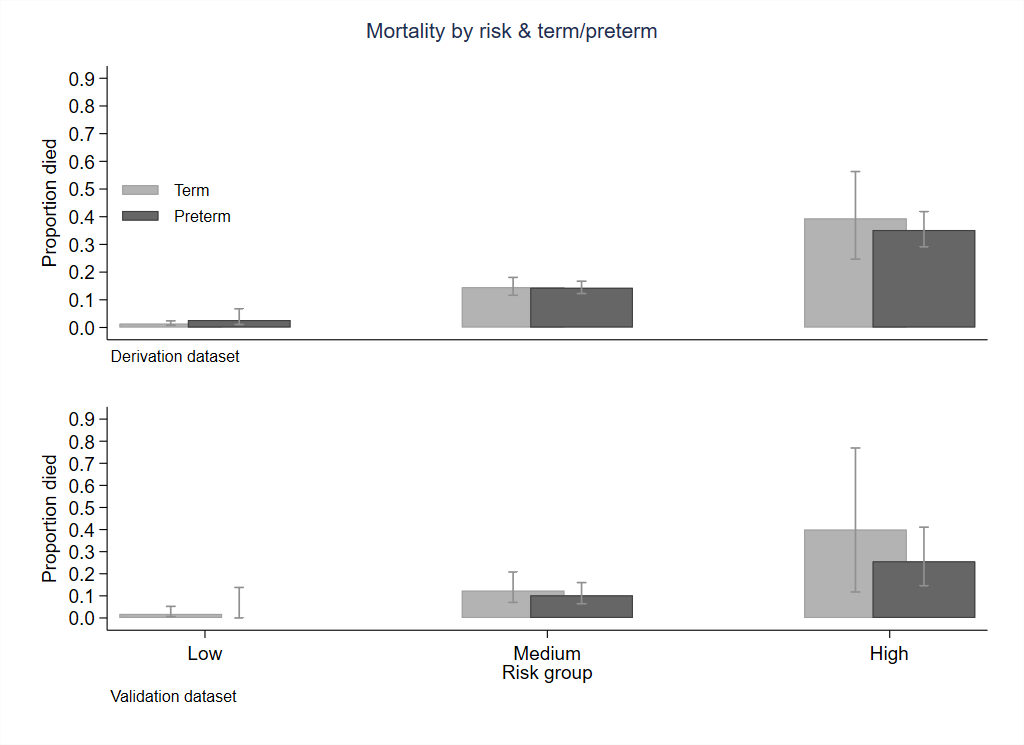

8. **Supplement figure 8: Comparison between WHO pSBI signs and NeoSep Severity Score for predicting 28-day mortality**

| **Score based on WHO signs of Clinical Severe Infection** | | **Modified NeoSep Severity Score** | |
| --- | --- | --- | --- |
| Derivation data | Validation data | Derivation data | Validation data |
| 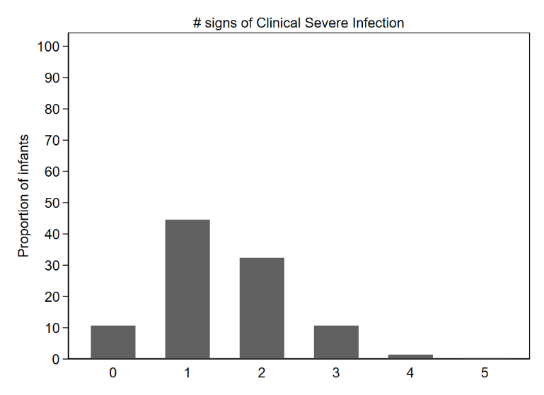 | 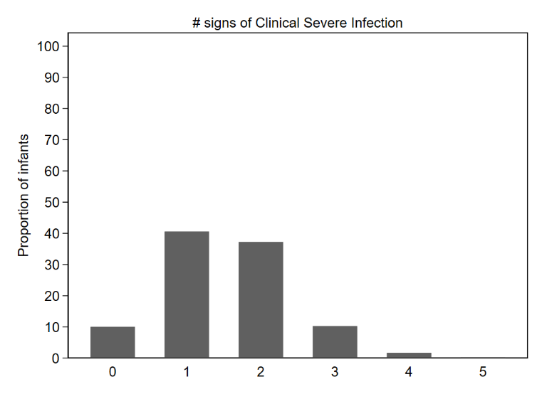 | 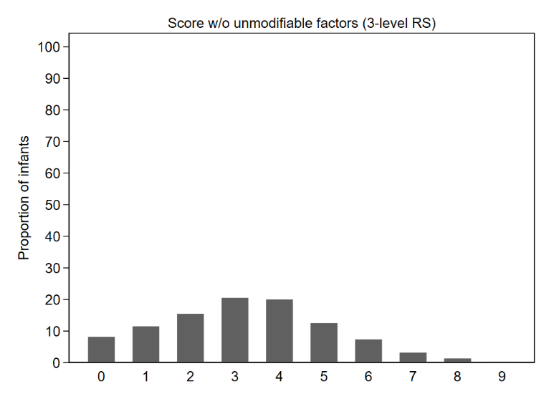 | 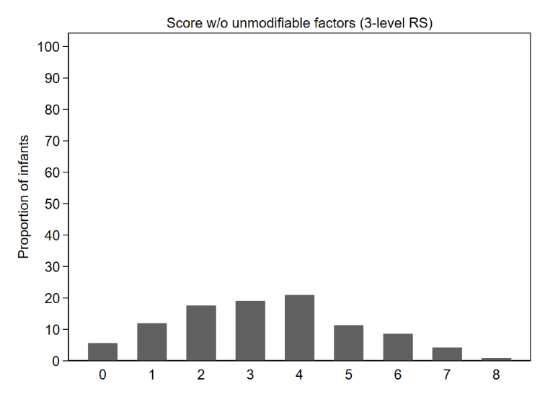 |
| 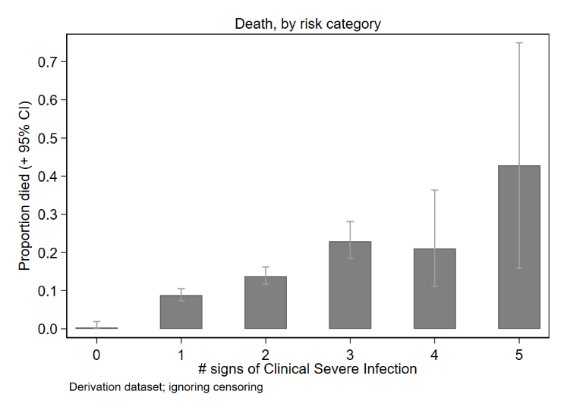 | 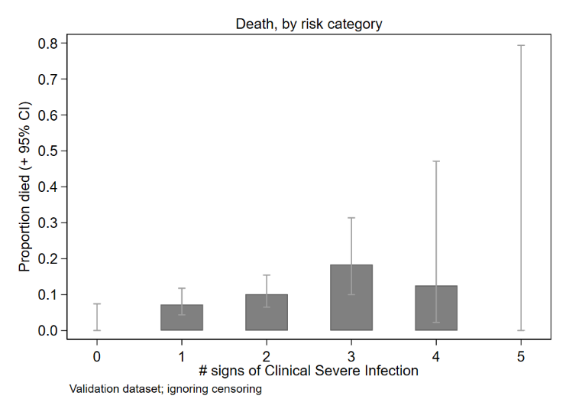 | 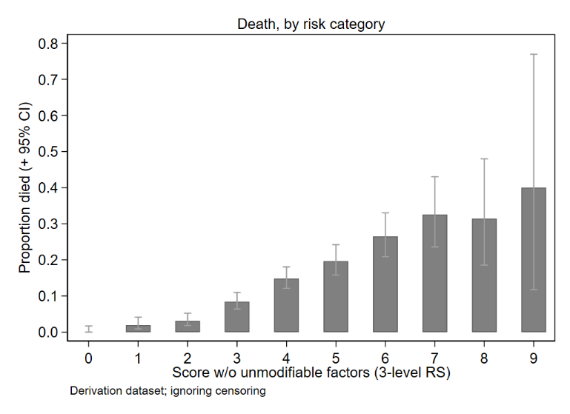 | 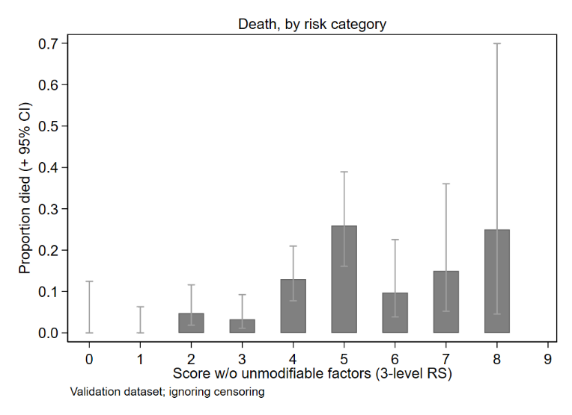 |
| Harrell C: 0.649 (0.621-0.675) | Harrell C: 0.629 (95% CI 0.559-0.699) | Harrell C: 0.741 (0.717-0.765) | Harrell C: 0.716 (0.651-0.774) |

Note: Score based on WHO signs of Clinical Severe Infection, compared with a modified NeoSep Severity Score excluding unmodifiable infant/birth characteristics.

9. **Supplement figure 9: NeoSep Recovery score on day 2 and change in score from baseline to day 2.**

**Derivation data**

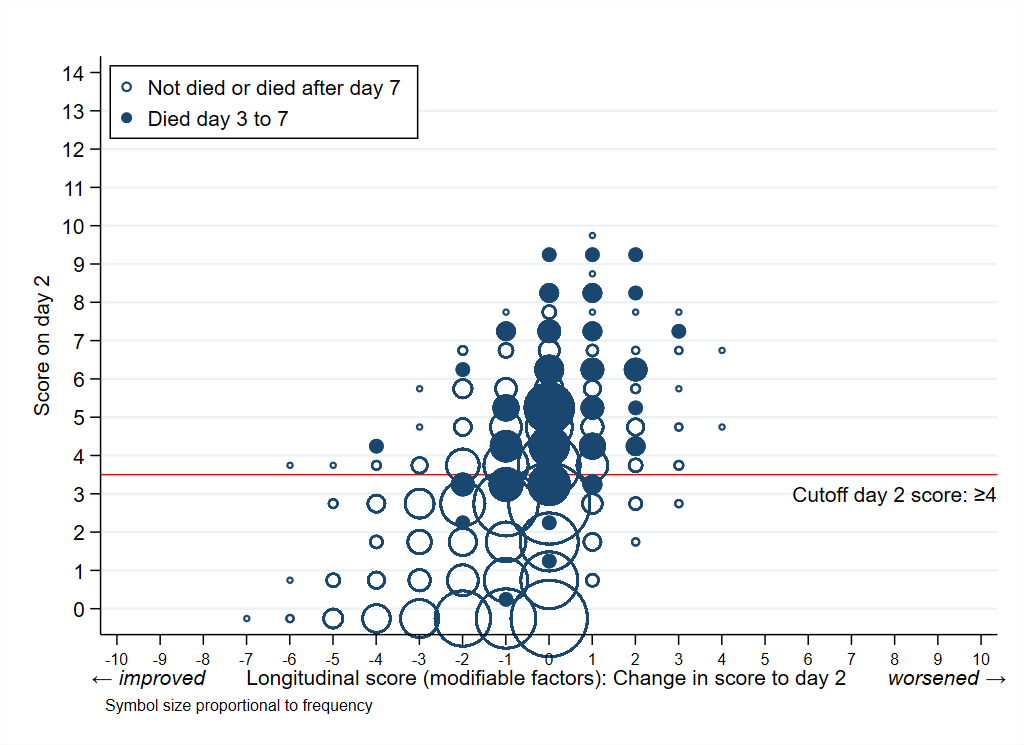

**Validation data**

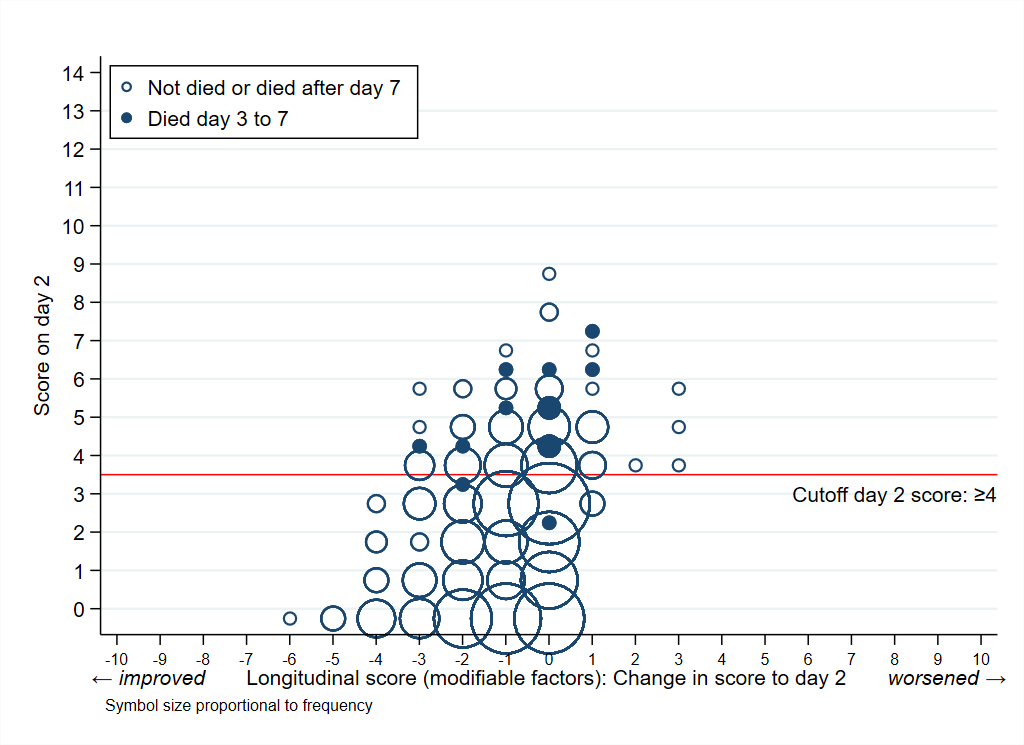

10. **Supplement table 1: Place of enrolment, time in hospital and age at admission**

|  | **N=3204** |
| --- | --- |
| What type of unit is the baby on at enrolment? |  |
| Level 1 NICU | 412 (12.9%) |
| Level 2 NICU | 584 (18.2%) |
| Level 3 NICU | 1377 (43.0%) |
| General Neonatal Medical Ward | 386 (12.0%) |
| Emergency Department | 2 (0.1%) |
| Paediatric Ward | 403 (12.6%) |
| PICU | 15 (0.5%) |
| Other | 25 (0.8%) |
| Time in hospital since admission (hrs), median (IQR) | 22 (1, 126) |
| Time in hospital (uninterrupted) |  |
| ≤48 hrs | 1886 (58.9%) |
| >48 hrs | 1318 (41.1%) |
| Age group |  |
| Neonate (<28 days) at admission | 3031 (94.6%) |
| Infant (28-x days) at admission | 173 (5.4%) |

12. **Supplement table 2: Comparison of factors in NeoOBS sepsis severity score with existing scores and sepsis criteria**

| **Mortality prediction, sepsis criteria & sepsis severity scores in neonates** | | | | | | | | | | |
| --- | --- | --- | --- | --- | --- | --- | --- | --- | --- | --- |
|  | | **General mortality prediction scores** | | | | **Existing Sepsis Criteria** | | | **Sepsis severity scoring** | |
|  | | **High income example** | **LMIC based** | | |  |  |  | **nSOFA sepsis severity score**^1,2^ | **NeoSep Sepsis**  **severity & recovery scores** |
|  |  | **CRIB II mortality prediction score**  *(& CRIB)* | **NMR 2000**^3^ | **SAWS score**^4^ | **Modified sick neonate score**^5^ | **pSBI criteria**^6^  *(*********=critical illness sign)* | **EMA**^7^ | **NeoOBS inclusion criteria** |  |  |
| **Purpose of score/criteria** | | Mortality prediction in LBW neonates | Mortality prediction in LBW neonates | Mortality prediction in LBW neonates | Mortality prediction in neonates | Clinical diagnosis of sepsis | Enrolment in clinical trials for sepsis | Enrolment in observational study of sepsis | Severity of organ failure and mortality prediction in sepsis |  |
| **Number of neonates from LMIC context included to generate score** | | - | 550  *(110,176 from high income)* | 428 | 585 | - | - | - | None  *(679 from high income)* | 3204 |
| **Relevant age for use of score** | | <12 hrs | <24hrs | <72hrs | Majority  <2 days | <60 days | <28 days | <60 days | 4-120 days  (95% <60d) | <60 days |
| **Weight category of included babies** | | LBW | <2kg | <1.5kg | Majority <2kg | >1.5kg | Any | Any | Majority <1.5kg (88%) | Any |
| **Specifically developed for sepsis?** | | No | No | No | No | Yes | Yes | Yes | Yes | Yes |
| **Adapted to LMIC context** | | No | Yes | Yes | Yes | Yes | No | Yes | No | Yes |
| **Predominantly community or hospital based** | | Hosp | Hosp | Hosp | Hosp | Comm | Hosp | Hosp | Hosp | Hosp |
| **Factors in score / criteria** | **Baby characteristics** | Birth weight  Sex  *Congenital malformation* | Gestation  Weight  Sex | Sex  Age  Weight  Gestation | Birth weight  Gestation | - | - | - | - | Birth weight  Time in Hospital  Gestational age  Congenital anomalies |
|  | **Vital signs** | Temp | SpO2 at admission | - | HR  SpO2  Glucose  CRT  Temp | Temp <35.5C  ≥38C | Temp <35.5C  ≥38C  Temp instability  HR<10^th^ centile  HR >2SD above normal | Temp  <35.5C  ≥38C  Temp instability  HR >180  HR <100 | - | Abnormal temp  (<35.5,  ≥38 – <39 °C, ≥39°C) |
|  | **Respiratory signs** | Min/Max FiO2 <12hrs | Max respiratory support | - | Increased respiratory effort | Fast breathing    Severe indrawing | Apnoea  Tachypnoea  Increased O2 requirement  Ventilatory support | Apnoea  Grunting  Cyanosis  Severe chest indrawing  Increased O2 requirement  Ventilatory support | SpO2/FiO2 ratio  Requirement for Invasive ventilation | O2 or ventilation support  Cyanosis (recovery score only) |
|  | **Circulatory signs** | - | - | - | - | - | Bradycardia    Tachycardia    Rhythm instability    Reduced urine output    Hypotension  Mottled skin  Impaired perfusion | CRT >3  or  Mottled skin  or  Other evidence of shock | Requirement for inotropes  Requirement for steroids for hypotension | Evidence of shock  (including any of: CRT >3secs, mottled skin, cold peripheries or ‘other’) |
|  | **Gastrointestinal signs** | - | - | - | - | Poor feeding  *Not feeding at all******** | Feeding intolerance  Poor sucking  Abdominal distension | Difficulty feeding  Or  Feeding intolerance  Abdominal distension | - | Difficulty feeding/ feeding intolerance  Abdominal distension |
|  | **Neurological signs** | - | - | - | - | Movement only when stimulated  Or  *no movement at all**    *Convulsions** | Irritability    Lethargy  Hypotonia | No movement  or  Movement only when stimulated  Bulging fontanelle  Lethargy or drowsiness  Hypotonia / floppiness  Abnormal posturing  Convulsions  Irritability | - | Lethargy, Reduced or no movement |
|  | **Skin / umbilicus** | - | - | - | - | - | Petechial rash  Sclerema | Petechial rash  Multiple or severe skin pustules  Pus from umbilical stump | - |  |
|  | **Laboratory signs** | Max base deficit <12h | - | - | - | - | WBC  <4 x10⁹  >20 x10⁹ cells/mL  IT ratio >0.2  Platelets <100 x10⁹ cells/mL  CRP >15mg/L  Procalcitonin ≥2 ng/ml  Glucose intolerance  Metabolic acidosis (BE <-10 mEq/L, Lactate >2mMol/L) | WBC  <4 x10⁹  >20 x10⁹ cells/mL  Neutrophils <1.5x10⁹ cells/mL  IT ratio >0.2  CRP >10mg/L  Metabolic acidosis  (BE <-10 mEq/L, Lactate >2mMol/L) | Severity of thrombocyto-paenia | - |

13. **Supplementary statistical methods**

We constructed a Cox regression model for 28-day mortality based on baseline clinical parameters known at sepsis presentation (i.e., before availability of microbiological results) and used this to develop a neonatal sepsis severity score. Baseline was defined as up to 24 hours from the time blood cultures were taken. Candidate predictors selected for initial consideration had missing values in <10% of the included infants, and had been found to be predictive in other studies^8^ or were *a priori* determined to be clinically important and not highly correlated with other factors (table 1). Factors that are not usually available in low-income settings including laboratory results (missing in >10%), and clinical signs with prevalence <5% were not considered. All analyses were based on available data, and imputation methods were not used. A 15% randomly selected sample per site was reserved for model validation and not used in any model development. This validation procedure was pre-defined because of expected large differences between sites in mortality, instead of randomly choosing one or two sites for validation which then might have turned out to be unrepresentative. In the remaining 85% of infants, model development used backwards elimination (exit p=0.05) to identify independent predictors of mortality, considering both categorical (e.g. presence of sepsis signs) and continuous predictors (birth weight, gestational age, time in hospital, age at baseline, oxygen saturation, respiratory rate, heart rate, temperature), taking into account the potential for non-linear relationships (e.g. where both high and low values are associated with poor outcomes) by using fractional polynomials. Fractional polynomials (FP) were implemented using the mfp command in Stata with power (-2,-1,-0.5,0,0.5,1,2) and significance level of 0.05 for testing between FP models of different degrees. Initial variable selection was done on complete cases which comprised 2313/2726 (85%) infants in the training set; final models were re-fitted to complete cases for the included factors (2705/2726 (99%) infants in the derivation sample). Interactions between birth weight and clinical predictors were examined and one strong interaction (p<0.001) was found with ventilation support; however, this was not included in the final model in favour of simplicity for use of the prediction model in clinical practice, and because including additional points for the interaction made very little difference to the C-statistic. The proportional hazards assumption was checked based on Schoenfeld residuals, and no deviations were found (p>0.05). A points-based risk score, where each predictor of death is assigned a number of points, was then developed from model coefficients: for each of the categorical factors, coefficients were divided by the smallest of the coefficients (lethargy) and rounded to the nearest integer. For the continuous factors, a clinically relevant reference value was chosen (birth weight: 3000g; temperature: 37°C; gestational age: 39 weeks; time in hospital: 14 days), and the number of points associated with lower/higher values was then based on the difference to the reference. This initial score was then further simplified to provide a more feasible and pragmatic scale for use in LMIC by assigning 1 score point for a predictor if its value in the initial score was <3, 2 score points if its initial value was 3-5 points, and 3 score points if its initial value was ≥6 points. For each participant, points were added with a higher score indicating a higher risk of death. Harrell’s C-index with bootstrapped confidence intervals was used as a measure of discrimination of the prognostic Cox models, and the Hosmer-Lemeshow goodness of fit test was applied after running a logistic model with death as the independent and the score as the dependent variable as a measure of calibration.

We compared the NeoSep Severity Score with a score based on WHO pSBI^6^, allocating one point for each of the following 6 signs: fever (≥38°C), low body temperature (<35°C), movement only on stimulation, feeding poorly, fast breathing (≥60 breaths per minute on days 0-6?), and severe chest indrawing. Whereas the first 5 signs were directly reported/measured in NeoOBS, the latter was only reported as part of a composite respiratory sign (severe chest wall in-drawing, increased oxygen requirement or need for ventilation); for the score calculation, one point was given if this was reported.

To estimate the association between daily risk of death after initiating IV antibiotics and time-updated factors, and hence develop a recovery score, we used Cox proportional hazards regression with time-varying independent factors. We started model building with all clinical predictors included in the baseline severity score as time-updated factors; unmodifiable infant and birth characteristics were excluded because they cannot evolve and would have restricted interpretation of the recovery score (e.g., pre-term babies would have a minimum score of 1 and could have never reached “full recovery” with a score of 0). Forward selection (entry p=0.05) was then used to identify any additional independent time-updated clinical predictors. Fractional polynomials were used for continuous factors as described above. For vital parameters, the last value was used per assessment period. To avoid selecting factors representing the mechanism of dying (e.g. stopping breathing = respiratory distress) rather than being predictors of subsequent death, clinical parameters reported on the previous day were used as predictors of death on the current day in all time-updated models. A points-based risk score was then derived similarly to the baseline severity score as described above. Discrimination was assessed using time updated Area Under the Receiver Operator Curves (AUROCs) ignoring censoring as this was low (<2% overall).

14. **References**

6.	WHO. *Managing possible serious bacterial infection in young infants when referral is not feasible*. (2015).

7.	Tuzun, F. *et al.* Is European Medicines Agency (EMA) sepsis criteria accurate for neonatal sepsis diagnosis or do we need new criteria? *PLoS One* **14**, e0218002–e0218002 (2019).

8.	Liang, L. *et al.* Predictors of Mortality in Neonates and Infants Hospitalized With Sepsis or Serious Infections in Developing Countries: A Systematic Review	. *Frontiers in Pediatrics*	**6**, 277 (2018).
